# Intensity-Driven Shifts in Tonotopic Coding in Humans: A Framework for Cochlear Implant Frequency Allocation

**DOI:** 10.1101/2025.05.05.25327002

**Authors:** Amit Walia, Matthew A. Shew, Shannon M. Lefler, Amanda J. Ortmann, Patrick Ioerger, Matthew Wu, Jordan Varghese, Jacques A. Herzog, Craig A. Buchman

**Affiliations:** Washington University School of Medicine, Department of Otolaryngology Head & Neck Surgery, 660 S. Euclid Ave. Campus Box 8115, St. Louis, MO 63110; University Hospitals Cleveland Medical Center, Cleveland, OH, USA, Department of Otolaryngology-Head and Neck Surgery, Case Western Reserve University School of Medicine, Cleveland, OH, USA

## Abstract

Tonotopic cochlear organization underlies auditory perception, yet cochlear implant (CI) programming typically employs fixed frequency-place maps not based on human physiology. Animal studies suggest intensity-dependent shifts in cochlear tuning, but this has not been confirmed in humans. Here, we demonstrate that cochlear tonotopy dynamically shifts basally with increasing sound intensity in humans. Using intracochlear electrocochleography from a 22-electrode array, we found that high-intensity stimuli (>80 dB SPL) shifted best-frequency locations basally by up to 158° (∼one octave) and significantly broadened cochlear excitation compared to threshold stimulation. This intensity-driven shift challenges static CI frequency mapping and supports a dynamic, intensity-adjusted approach that better replicates natural cochlear processing. Implementing such intensity-based frequency allocation in cochlear implants may reduce place-frequency mismatch, potentially enhancing critical auditory outcomes for CI users, including speech recognition in complex listening environments and improved music perception.

**Teaser:** Higher sound intensity shifts the ear’s internal frequency map, revealing dynamic hearing mechanics in humans.

## Introduction

Sound pressure waves enter the ear canal and set off a chain of vibrations within the cochlea, resulting in the displacement of the basilar membrane. High-frequency sounds peak at the basal end of the cochlea, while low-frequency sounds peak at the apical end (*1*). Outer hair cells, when stimulated at low intensities, amplify the basilar membrane vibrations, creating a sharply-tuned and localized mechanical response. Inner hair cells at the same location transduce these vibrations and send signals to the auditory nerve fibers. Each auditory nerve has a characteristic frequency (CF) that matches its tonotopic location along the cochlea, reflecting the basilar membrane’s mechanical response. Numerous studies across mammalian species support this concept, showing nearly identical frequency responses between basilar membrane vibrations and auditory neuron responses (*2–11*).

The intensity of a sound stimulus can also impact the location of maximal basilar membrane vibration within the cochlea. Early research on temporary threshold shifts revealed that intense sounds could cause a basalward shift in the basilar membrane traveling wave, moving the peak frequency response by roughly half an octave (*12–14*). Later studies in guinea pigs supported this phenomenon; cochlear microphonic (CM) recordings indicated that higher sound levels led to shifts in sensitivity and voltage peaks toward the base of the cochlea. (*15*). For example, Russell and Nilsen (1997) found that as sound intensity increased above 60 dB SPL, the peak basilar membrane response for 15 kHz tones shifted about 0.5 mm toward the base, demonstrating how higher intensity spreads and shifts the basilar membrane’s response (*16*).

Greenwood was the first to develop a frequency-position equation to estimate CF along the organ of Corti (*17, 18*). His function was based on human critical bandwidths derived from masked threshold audiometry, which he theorized followed an exponential relationship with distance along the cochlear partition, corresponding to a consistent physical distance on the basilar membrane. Although the spiral ganglion (SG) and organ of Corti (OC) spiral together along the modiolar axis, they differ significantly: the SG is about 40% shorter, has fewer rotations (approximately 2 compared to the OC’s 2¾ turns), and the radial nerve fibers take a more skewed path from the SG to the OC, particularly toward the apex. Consequently, Greenwood’s frequency-position estimates for the OC do not directly apply to the SG, even when adjusted for length or angle. Stakhovskaya et al. (2007) provided an anatomical correction by analyzing human temporal bones, tracing radial fibers from the OC to their corresponding SG locations to create a frequency-matched map(*19*) More recently, Li et al. (2021) utilized synchrotron radiation phase-contrast imaging to generate a 3D reconstruction of the basilar membrane and SG positions, producing a frequency-position function for the SG that mirrors Greenwood’s, but with different coefficient values.(*20*) Although the frequency-place maps developed by Stakhovskaya et al. (2007) (*19*) and Li et al. (2021) (*20*) differ slightly, particularly near the apex, they remain within 0.5% of each other in terms of SG position relative to frequency, representing the current gold standard for evaluating cochlear frequency-position relationships, particularly for cochlear implant mapping. A significant limitation of these maps is that both Greenwood’s function (1961) (*18*) and Stakhovskaya’s application (*19*) are derived from frequency representations at threshold-level stimulation in individuals with normal hearing.

Our group has recently focused on utilizing intracochlear electrocochleography (ECochG) during cochlear implantation to better understand *in vivo* place coding within the human cochlea. We conduct intracochlear ECochG by delivering pure tone stimuli through a sound delivery tube into the ear canal, capturing the resulting electrophysiological responses along the multi-electrode array. Our previous work has demonstrated variability in the ECochG map among subjects with hearing loss, likely due to anatomical differences within the cochlea among individuals (*21, 22*). When compared to the Greenwood OC map and the Stakhovskaya SG map, ECochG measures in two patients with preserved cochlear function suggested that stimulus intensity may be important. Therefore, a personalized electrophysiological mapping approach may provide a more accurate frequency-position function for cochlear implant programming. However, exploring the effects of decreasing stimulus intensity and observing apical shifts, as seen *in vivo* in guinea pig studies (*16*), is challenging in subjects with hearing loss because responses are typically not detectable at lower intensities.

Auditory neuropathy spectrum disorder (ANSD) presents a unique opportunity for studying cochlear function using intracochlear ECochG due to its distinct characteristics. Patients with ANSD maintain normal (or near normal) outer hair cell function, as evidenced by preserved otoacoustic emissions and cochlear microphonics, despite displaying abnormal or absent auditory brainstem responses, which indicate disruptions in auditory nerve transduction (*23, 24*). With intact outer hair cells, ECochG measurements in ANSD patients can accurately reflect outer hair cell tuning curves, offering valuable insights into *in vivo* place coding and frequency mapping within the cochlea. As a result, ANSD patients provide an excellent model for advancing our understanding of cochlear function, particularly in exploring intensity-dependent shifts.

The purpose of this study is to demonstrate that increasing intensity levels lead to broader frequency tuning and a basal shift in place coding within the human cochlea. Additionally, we aim to quantify the magnitude and direction of these shifts to develop a more accurate and dynamic tonotopic map benchmark for cochlear implants, moving beyond the limitations of a fixed frequency-position map and replicating the adaptive capabilities of the normal ear.

## Results

### Intensity-dependent shifts in best-frequency position

Figure 1 illustrates the detailed experimental setup used to investigate intracochlear ECochG responses to auditory stimuli. During cochlear implantation surgery, calibrated pure-tone stimuli were delivered via a foam ear-tip inserted into the external auditory canal, and electrophysiologic responses were simultaneously captured through a 22-electrode cochlear implant array positioned within the cochlea through a round window opening (Fig. 1A, B). Recorded waveforms were processed into difference curves, calculated by averaging condensation and rarefaction phase responses [(condensation-rarefaction)/2], and subsequently analyzed using fast Fourier transforms (FFT) to determine patient-specific tuning curves at different frequencies and stimulus intensities (Fig. 1C).

**Fig. 1.**
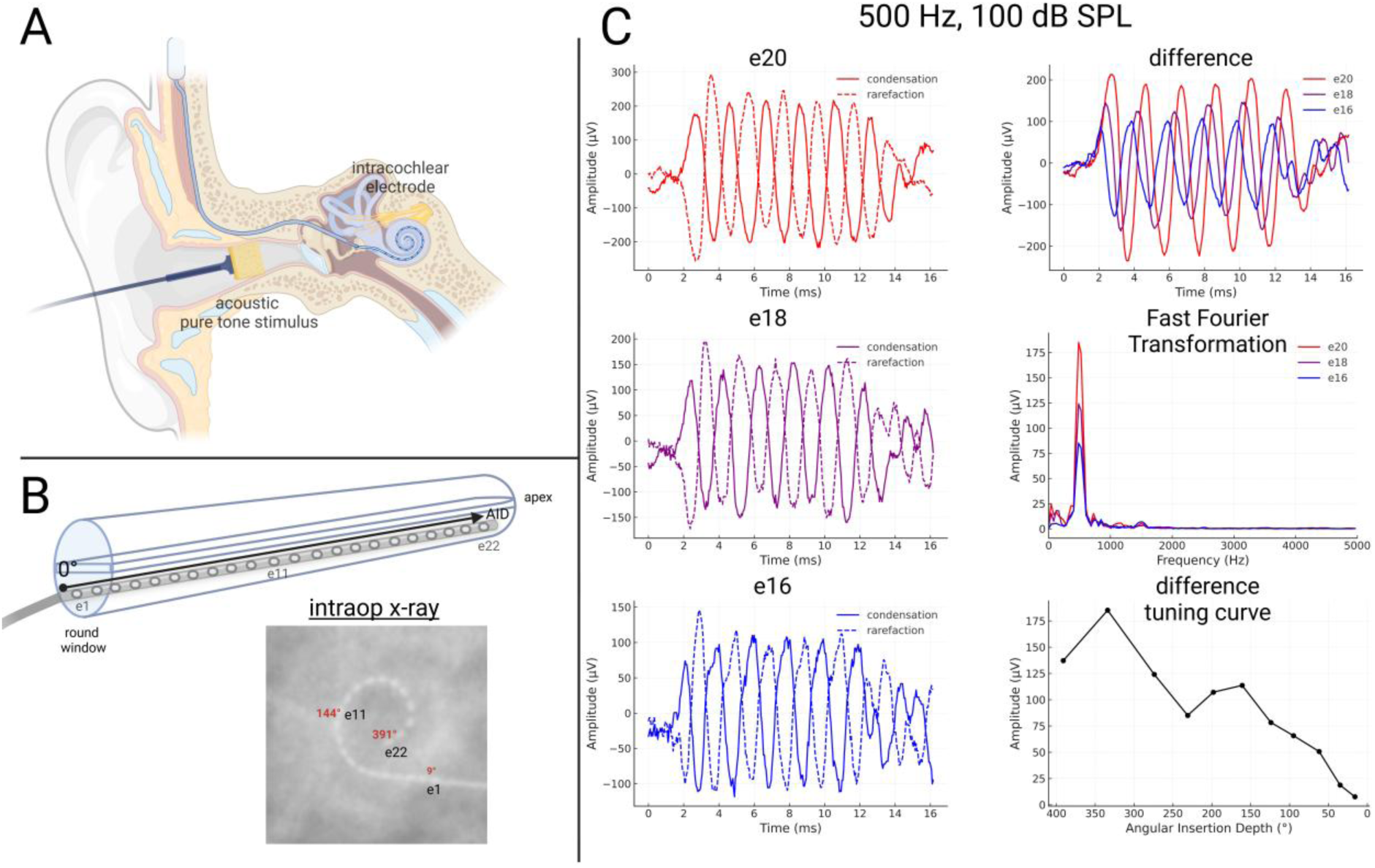
Intraoperative electrophysiologic recording setup and sample data. (**A**) During cochlear implant surgery, a foam ear-tip is placed in the external auditory canal (before sterile preparation) to deliver a pure-tone stimulus. Through a mastoidectomy and facial recess approach, the electrode array is inserted via a round window-related opening, allowing intracochlear recordings from each contact. Temporal bone model created with BioRender.com (**B)** An intraoperative X-ray confirms electrode placement. The round window (0°) serves as a reference for angular insertion depth. The array contains 22 electrode contacts (e1 = most basal, e22 = most apical). A validated helical scala tympani model is applied to the X-ray to determine each electrode’s position. (**C**) Example waveforms recorded for a 500 Hz, 100 dB SPL stimulus. Condensation and rarefaction phases are collected from multiple electrodes; the “difference” waveform is computed as ^𝑐𝑜<u>𝑛</u>𝑑𝑒<u>𝑛</u>𝑠𝑎𝑡<u>𝑖</u>𝑜𝑛<u>−</u>𝑟𝑎𝑟𝑒𝑓𝑎𝑐𝑡<u>𝑖</u>𝑜<u>𝑛</u>^. Fast Fourier transforms of these difference waveforms yield response amplitudes, which can be plotted as tuning curves for each frequency and intensity.

Clear intensity-dependent shifts in cochlear tonotopic mapping emerged from these analyses. Figure 2 presents representative CM tuning curves obtained from a subject with ANSD who demonstrated preserved outer hair cell function, confirmed through distortion product otoacoustic emissions. As stimulus intensity progressively increased from near-threshold levels (∼43 dB SPL) to above 80 dB SPL, the best-frequency (BF) electrode location systematically shifted basally (toward higher-frequency electrode positions), accompanied by a notably broader cochlear excitation pattern. Conversely, lower stimulus intensities elicited localized responses, with maximal activation confined to a few electrode electrodes, indicating a narrower and more spatially limited excitation profile (Fig. 2).

**Fig. 2.**
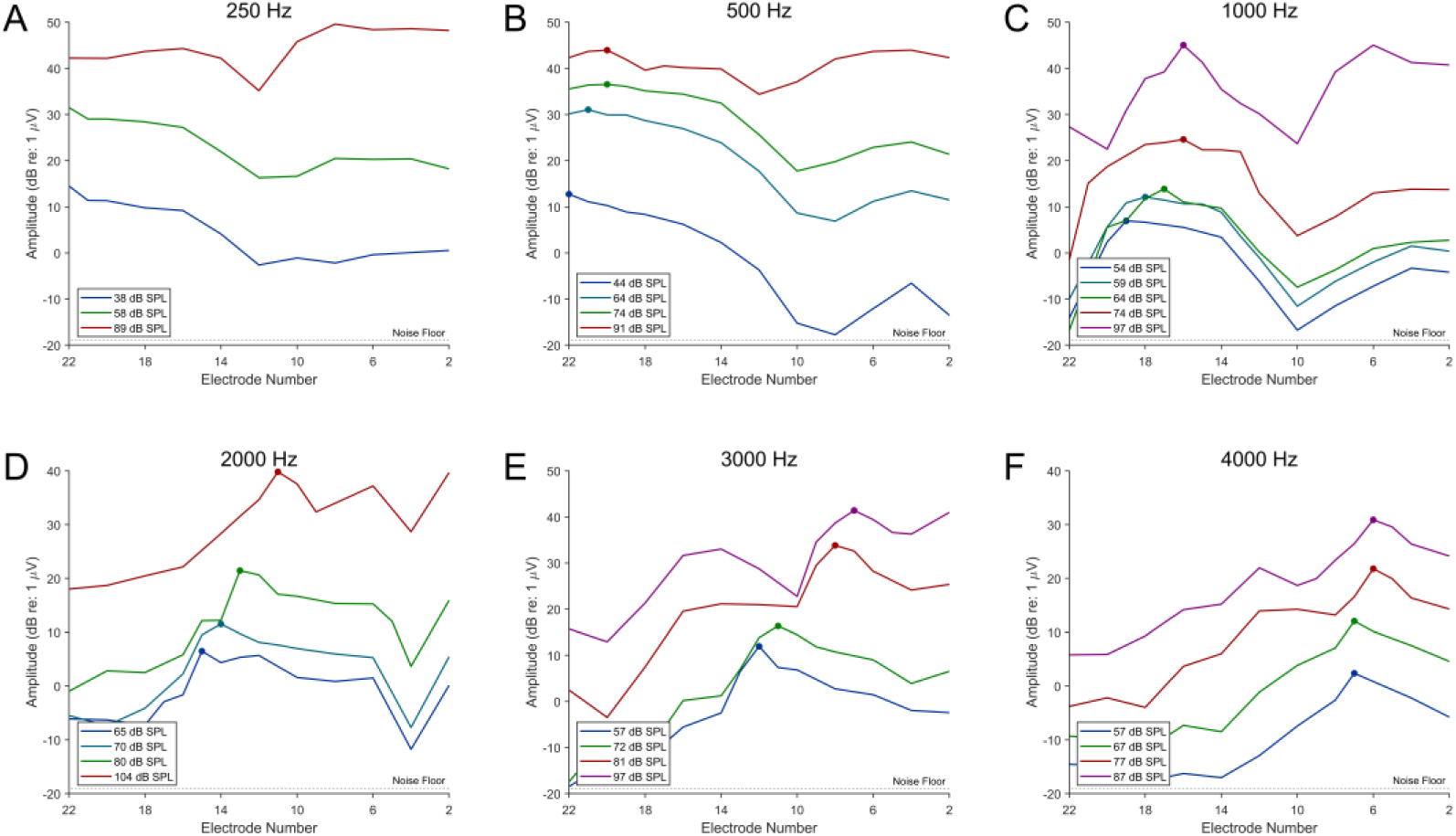
Cochlear microphonic tuning curves demonstrating intensity-dependent shifts. This Fig. illustrates example tuning curves from a subject with auditory neuropathy spectrum disorder, in whom preserved outer hair cell function is confirmed by distortion product otoacoustic emissions. The cochlear microphonic tuning curves demonstrate that as stimulus intensity increases from threshold, the electrode exhibiting the maximum response (i.e., the best frequency electrode) shifts basally, and the spectral spread of the acoustically evoked response broadens. Conversely, at lower intensities, the response becomes more spatially confined.

### Comparison of ECochG-derived maps with established frequency-position functions

To further quantify these intensity-dependent cochlear shifts, ECochG-derived best-frequency locations were systematically compared with established cochlear tonotopic maps, including Greenwood’s traditional threshold-based frequency-position map (*17*) and more recent maps from Stakhovskaya et al. (*19*) and Li et al. (*25*) (Fig. 3). At higher stimulus intensities (∼83 dB SPL), the ECochG-derived maps significantly deviated from Greenwood’s threshold-based predictions (Fig. 3A). Reductions in stimulus intensity produced progressive apical shifts in best-frequency positions (Fig. 3B**–E**), with statistical analyses confirming significant differences from Greenwood’s frequency-position map at all intensity levels evaluated (Wilcoxon signed-rank test, *p* < 0.05; Fig. 3C**–E**). At near-threshold intensities (∼43 dB SPL), the ECochG-derived BF map differed significantly from Greenwood’s predictions (Wilcoxon signed-rank test, *z* = 5.00, *p* < 0.001).

**Fig. 3.**
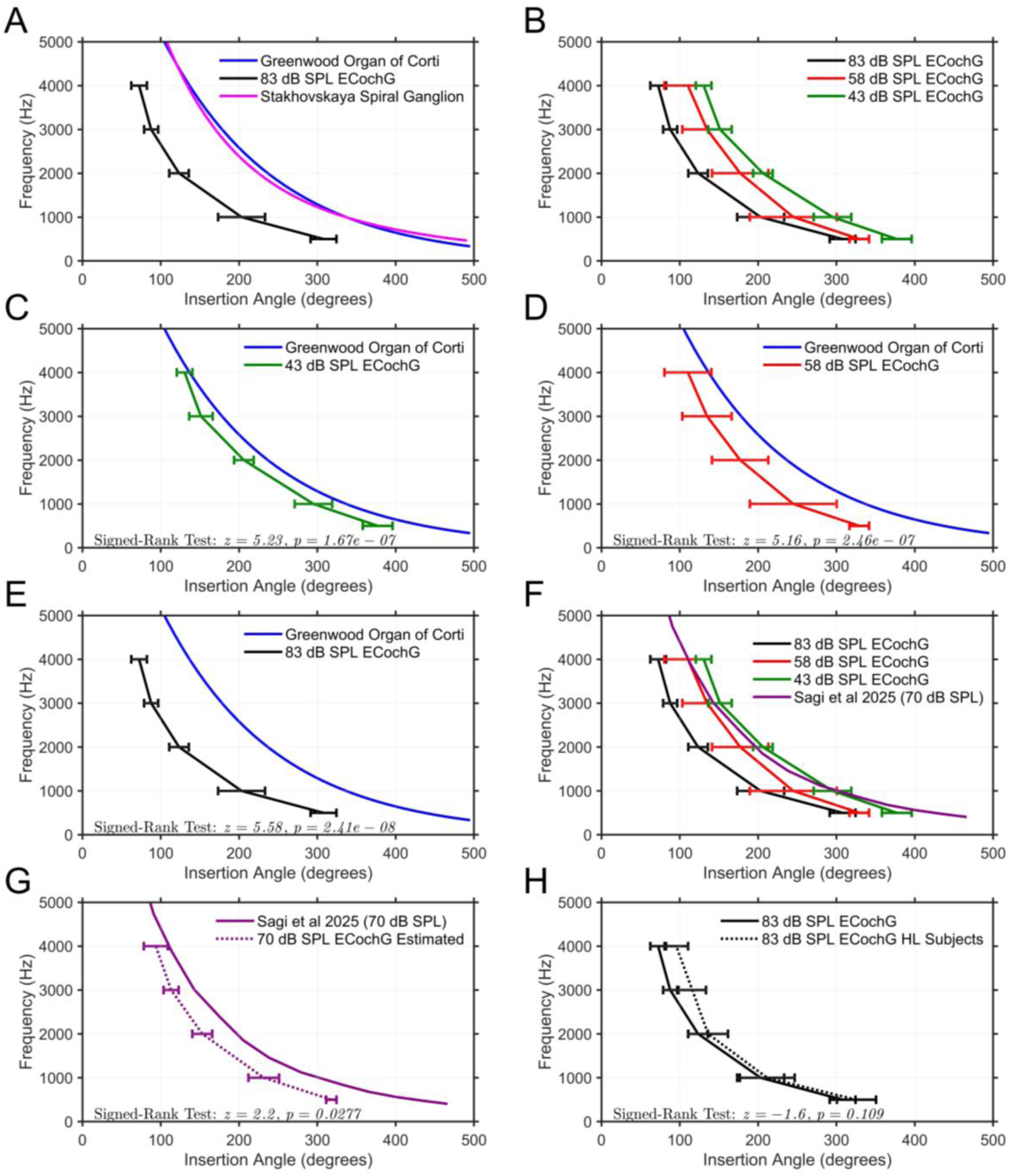
Comparison of ECochG-derived and established frequency-position maps. Panels (**A**)–(**H**) illustrate how cochlear tonotopy shifts with stimulus intensity and how these shifts compare to established frequency-position functions. **(A)** High-intensity (83 dB SPL) ECochG-derived map (black data) overlaid on the threshold-based Greenwood(*17*) (blue) and Stakhovskaya(*19*) (pink) functions. **(B)** ECochG-derived maps at three intensities (∼43, ∼58, and ∼83 dB SPL), demonstrating the effect of stimulus level on best-frequency (BF) location. **(C–E)** Comparisons of ECochG-derived maps at 43 dB SPL (C), 58 dB SPL (D), and 83 dB SPL (E) against Greenwood’s threshold-based function, with Wilcoxon signed-rank test results indicating significant differences (*p* < 0.05). **(F)** ECochG-derived maps at 43, 58, and 83 dB SPL compared with the intensity-adjusted map from Sagi et al. (2025), which extrapolates animal data obtained at ∼70 dB SPL. **(G)** The animal-based Sagi map at 70 dB SPL compared with the ECochG- derived 70 dB SPL map predicted from a mixed model. The predicted curve (with 95% confidence intervals) differed significantly from the Sagi map (Wilcoxon signed-rank test, *p* = 0.02). **(H)** Comparison of ECochG-derived maps at 83 dB SPL between subjects with auditory neuropathy spectrum disorder (ANSD; solid line, *N* = 8) and subjects with adult-onset post-lingual hearing loss(*21, 22*) (HL; dotted line, *N* = 50). The Wilcoxon signed-rank test indicated no significant difference between these two groups (*p* = 0.109). Error bars represent ±2 standard deviations. These comparisons highlight the variability in ECochG-derived maps at different intensities and emphasize the importance of incorporating stimulus level into frequency-position modeling.

At moderate stimulus intensities (∼70 dB SPL), ECochG-derived maps showed closer alignment with the recently proposed intensity-adjusted cochlear maps derived from animal models (Sagi et al., 2025 (*26*)); however, it was still significantly apically shifted compared to the ECochG map measured at the same intensity (Wilcoxon signed-rank test, *z* = 2.2, *p* = 0.028; Fig. 3G). Moreover, comparative analysis between ANSD subjects and previously studied adult-onset post-lingual hearing loss populations (*21, 22*) revealed no statistically significant differences in ECochG-derived maps at elevated stimulus intensities (∼83 dB SPL; Wilcoxon signed-rank test, z = 1.6, p = 0.109; Fig. 3H).

### Cochlear microphonic tuning curves in a subject with normal hearing

To validate the generalizability of these findings, cochlear microphonic tuning curves were also recorded from a subject with clinically normal hearing thresholds who underwent a translabyrinthine surgical procedure for acoustic neuroma removal (Fig. 4). Analysis of these data revealed similarly pronounced basal shifts in best-frequency electrode positions and broadening of cochlear excitation with increasing stimulus intensities. These findings closely paralleled the shifts observed in subjects with ANSD, reinforcing the interpretation that intensity-dependent cochlear frequency shifts are fundamental physiological responses rather than specific consequences related to cochlear pathology.

**Fig. 4.**
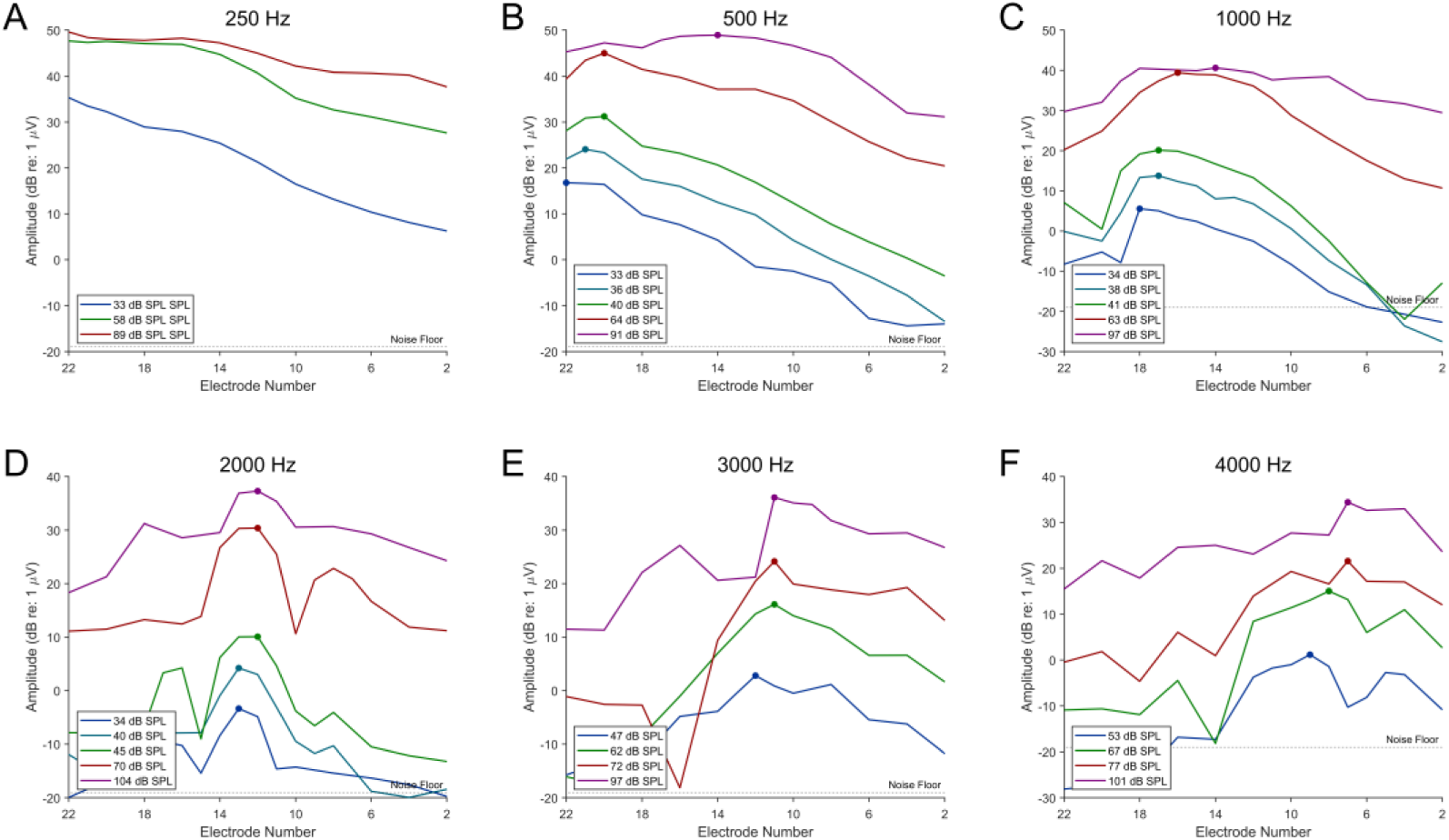
Cochlear microphonic tuning curves in a subject with normal hearing. This figure presents the CM tuning curves from a subject with normal pure-tone thresholds and word recognition scores who underwent a translabyrinthine craniotomy for vestibular schwannoma resection. The cochlear implant electrode was inserted prior to labyrinthectomy and removed after recording. As the acoustic stimulus intensity increased, the CM tuning curves shifted basally, with a broader and more diffuse spectral spread observed across the electrode array, similar to that seen for the subjects with ANSD.

### Quantification of intensity-dependent BF shifts across frequencies

Across all frequencies tested (500, 1000, 2000, 3000, and 4000 Hz), the AID of the BF electrode shifted systematically with changes in stimulus intensity for each individual tested (Fig. 5). Figure 5A displays the AID for BF electrodes across all frequencies. Figures 5B**–F** present log-mixed-effects fits for each frequency using the model:

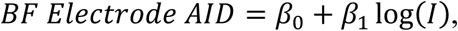

where 𝐼 is the stimulus intensity in dB SPL. At 500 Hz, the model indicated an intensity increase from near-threshold levels (∼44.0 dB SPL) to mid-intensity levels (∼79.0 dB SPL) was associated with a mean basal shift in AID of 45.3° (adjusted 𝑅^2^ = 0.821; *p* < 0.001). Comparable shifts were observed at 1000, 2000, 3000, and 4000 Hz, with mean shifts of 63.8°, 41.4°, 28.5°, and 39.4°, respectively. Corresponding adjusted 𝑅^2^ values were 0.828, 0.661, 0.788, and 0.650, with statistical analyses confirming these shifts were significant (*p* < 0.001 for 1000, 2000, and 3000 Hz; *p* = 0.029 for 4000 Hz). Across frequencies, the log transformation of intensity revealed a relationship that flattened at higher stimulus levels, suggesting diminishing sensitivity of the frequency–place function to intensity changes. Individual subject data across frequencies and intensities are summarized comprehensively in **Table 1**. Using these frequency-specific equations derived from log-mixed-effects fits, an estimated ECochG-based cochlear map at 70 dB SPL was generated (Fig. 3G). This ECochG-based map showed a significant basal shift compared to the animal-derived frequency-place map by Sagi et al. (*26*) at 70 dB SPL (signed-rank test: z = 2.2, *p* = 0.03).

**Fig. 5.**
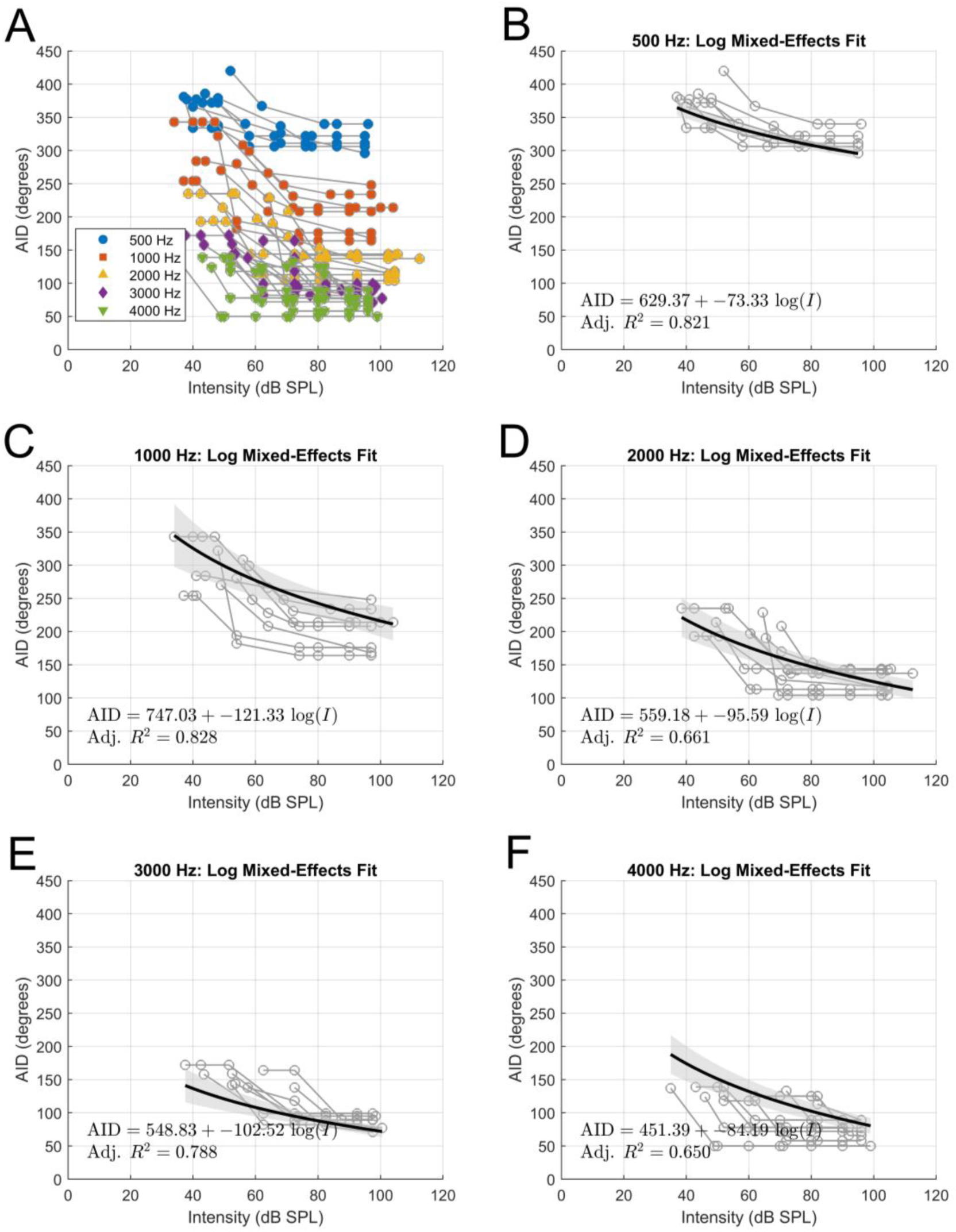
Best frequency electrode position as a function of stimulus intensity. Panel A displays combined data across all subjects for five frequencies (500, 1000, 2000, 3000, and 4000 Hz), with each subject’s BF electrode angular insertion depth (AID) plotted as connected gray lines and individual data points overlaid using distinct markers/colors for each frequency. Panels B–F present, for each specific frequency, the log-mixed-effects model fit relating stimulus intensity (dB SPL) to AID. In each panel, the best-fit curve is shown as a black line with a shaded 95% confidence interval, and the fitted equation with adjusted *R^2^* is annotated in the bottom left. All subplots share uniform axis limits (Intensity: 0–120 dB SPL; AID: 0–450°), facilitating direct comparison of intensity-dependent shifts in electrode position across frequencies.

**Table 1.** Individual subject data of stimulus intensity and best-frequency electrode shifts. Stimulus intensity levels (high, mid, and low) for each tested frequency (500, 1000, 2000, 3000, and 4000 Hz) across individual subjects. Corresponding angular insertion depths (AID) of the best-frequency (BF) electrodes and the magnitude of basal shifts (in degrees) between intensities are summarized. Data presented as mean (± SD) across subjects.

| Frequency (Hz) |  | Subject 1 | Subject 2 | Subject 3 | Subject 4 | Subject 5 | Subject 6 | Subject 7 | Subject 8 | Mean (SD) AIDBF (°) at Mean (SD) Low Intensity (dB SPL) | Mean (SD) Basal Shift* at Mean (SD) Stim level (dB SPL) | Mean (SD) Basal Shift** at Mean (SD) Stim level (dB SPL) |
| --- | --- | --- | --- | --- | --- | --- | --- | --- | --- | --- | --- | --- |
| 500 Hz | High Intensity (dB SPL), AID Basal Shift (°) | 101.5,80 | 100.5,66 | 100.5,70 | 100.5,73 | 100.5,50 | 100.5,75 | N/A | 100.5,117 | 383 (18.1) | 75.9 (20.5), 100.6 (0.4) | 50.6 (15.2), 63.4 (8.84) |
|  | Mid Intensity (dB SPL), AID Basal Shift (°) | 67.5,53 | 53.5,39 | 73.5,40 | 73.5,42 | 63.5,50 | 51.5,47 |  | 60.5,83 |  |  |  |
|  | Low Intensity (dB SPL), AID (°) | 51,420 | 43,373 | 39,366 | 47,379 | 42,372 | 36,381 |  | 44,391 |  |  |  |
| 1000 Hz | High Intensity (dB SPL), AID Basal Shift (°) | 104.0,65 | 104.0,36 | 104.0,102 | 104.0,72 | 104.0,158 | 104.0,78 | 111.0,129 | 99.0,94 | 295 (29.0) | 91.8 (38.4), 104.3 (3.2) | 76.6 (37.6), 71.1 (7.1) |
|  | Mid Intensity (dB SPL), AID Basal Shift (°) | 76.0,51 | 71.0,18 | 71.0,62 | 71.0,52 | 61.0,140 | 61.0,60 | 79.0,129 | 79.0,77 |  |  |  |
|  | Low Intensity (dB SPL), AID (°) | 58,299 | 41,284 | 49,270 | 54,280 | 48,322 | 37,254 | 34,343 | 56,308 |  |  |  |
| 2000 Hz | High Intensity (dB SPL), AID Basal Shift (°) | 111.0,55 | 111.0,77 | 111.0,95 | 111.0,72 | 111.0,125 | 111.0,81 | 119.0,98 | 112.0,70 | 207.5 (17.2) | 84.1 (21.5), 112.1 (2.8) | 75.2 (26.8), 78.3 (2.1) |
|  | Mid Intensity (dB SPL), AID Basal Shift (°) | 79.0,55 | 77.0,66 | 78.0,70 | 81.0,37 | 79.0,125 | 74.0,81 | 80.0,98 | 79.0,70 |  |  |  |
|  | Low Intensity (dB SPL), AID (°) | 58,197 | 40,193 | 68,208 | 63,190 | 62,229 | 44,194 | 36,235 | 37,214 |  |  |  |
| 3000 Hz | High Intensity (dB SPL), AID Basal Shift (°) | 100.0,60 | 105.0,70 | 105.0,39 | 105.0,67 | 105.0,74 | 105.0,59 | 108.0,95 | 106.0,49 | 193.4 (18.7) | 64.1 (16.9), 104.9 (2.2) | 56.2 (19.7), 76.5 (5.6) |
|  | Mid Intensity (dB SPL), AID Basal Shift (°) | 70.0,60 | 80.0,70 | 82.0,39 | 80.0,20 | 84.0,74 | 70.0,59 | 73.0,79 | 73.0,49 |  |  |  |
|  | Low Intensity (dB SPL), AID (°) | 52,142 | 52,159 | 72,138 | 57,138 | 62,164 | 43,158 | 37,172 | 53,144 |  |  |  |
| 4000 Hz | High Intensity (dB SPL), AID Basal Shift (°) | 105.5,59 | 105.5,55 | 105.5,24 | 105.5,60 | 105.5,56 | 105.5,50 | 108.5,87 | 101.5,46 | 127.0 (9.0) | 59.0 (13.3), 105.4 (1.9) | 54.6 (17.5), 78.1 (8.0) |
|  | Mid Intensity (dB SPL), AID Basal Shift (°) | 92.0,59 | 81.5,55 | N/A | 71.5,60 | 81.5,56 | 71.5,50 | 79.5,87 | 69.5,46 |  |  |  |
|  | Low Intensity (dB SPL), AID (°) | 59,125 | 55,127 | 48,113 | 55,118 | 58,133 | 46,139 | 38,137 | 49,124 |  |  |  |

\*High Intensity - Low Intensity (°)
\*\*Mid Intensity - Low Intensity (°)

### Intensity-dependent changes in spatial excitation spread

The spatial distribution of the acoustically-evoked responses was evaluated as a function of stimulus intensity and is shown in Fig. 6. At high-intensity stimulation levels (>80 dB SPL) across 500 Hz to 4 kHz, responses were broadly distributed across the entire electrode array, with the maximum amplitude consistently recorded at the electrode corresponding to the BF electrode (by definition). In contrast, as stimulus intensity decreased below 80 dB SPL, the activation became increasingly localized. Near threshold levels, only 1–3 electrodes exhibited measurable responses, indicating a more confined distribution of acoustically-evoked electrical activity. To quantitatively assess this effect, a linear mixed-effects model was fit to the outcome measure (i.e., the number of activated electrodes) for a 500 Hz stimulus, with “Intensity” and “IntensitySquared” included as fixed predictors and a random intercept by subject. The model revealed a significant positive linear effect of intensity (β = 0.8982, SE = 0.2266, t(33) = 3.964, *p* = 0.00037, 95% CI [0.437, 1.359]), confirming that the number of activated electrodes increased with increasing stimulus intensity. In addition, the quadratic term was significantly negative (β = −0.00555, SE = 0.00183, t(33) = −3.026, *p* = 0.00477, 95% CI [−0.00928, −0.00182]), implying that the increase in activation tapered off at higher intensities, likely due to the limited number of electrodes on the device. These results demonstrate that as stimulus intensity increases, the spread of the response increases in a nonlinear fashion. This relationship was similar across other tested frequencies 1 kHz – 4 kHz (Fig. 6B**-E**). Also, spread of excitation appeared greater at lower intensities for low frequency stimuli when compared to high frequency stimuli.

**Fig. 6.**
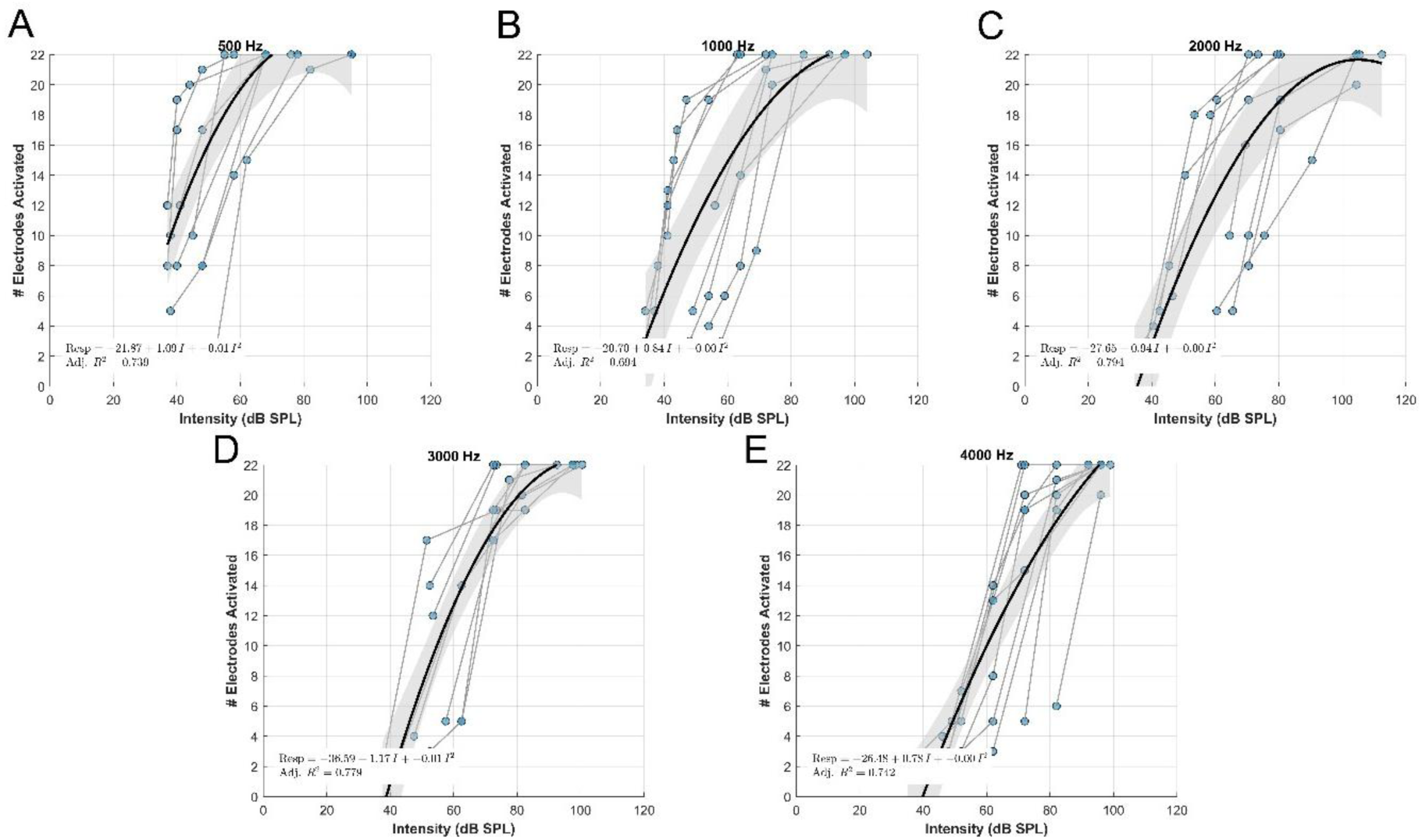
Spatial spread of excitation as a function of stimulus intensity. Each panel (**A–E**) shows the number of activated electrodes (y-axis, up to 22) as a function of stimulus intensity (x-axis, 0–120 dB SPL)) for a single frequency: 500, 1000, 2000, 3000, or 4000 Hz. Colored markers denote the predicted intensity at which the model forecasts 25%, 50%, 75%, and 100% electrode activation. Observed data from multiple subjects are displayed as scatter points with subject-specific lines, and a quadratic mixed-effects fit (solid curve) is overlaid with a 95% confidence band (shaded area). At lower intensities, fewer electrodes are activated, whereas high intensities elicit a broader spread of excitation that plateaus once all 22 electrodes are activated.

## Discussion

This study provides direct electrophysiological evidence that sound intensity significantly influences cochlear frequency-place coding in humans, revealing systematic basalward shifts in best-frequency (BF) response locations as stimulus intensity increases. These findings align with established animal model data demonstrating similar shifts driven by intensity-dependent cochlear mechanics (*16*). At lower sound intensities, outer hair cells (OHCs) enhance frequency selectivity and sensitivity through nonlinear amplification of basilar membrane motion, a process known as the “cochlear amplifier” (*27, 28*). Conversely, higher intensity stimuli saturate these active mechanisms, and passive mechanics dominate, producing broader traveling wave responses and basalward shifts in frequency-place coding (*16*).

Although these mechanisms have been extensively characterized in animal models, the precise nature of intensity-dependent shifts in the human cochlea has remained unclear. Our study addresses this gap by quantifying both the extent and spatial distribution of cochlear responses to a broad range of stimulus intensities—from near-threshold (33 dB SPL) to very loud levels (103 dB SPL). Figure 3, for example, illustrates individual electrode insertion angles relative to the place-frequency map predicted by Greenwood’s characteristic frequency (CF) function, alongside ECochG-derived maps at various intensities. The observed variability among subjects, even at similar intensity levels, underscores the role of inherent anatomical and physiological differences in shaping these responses.

Using intracochlear electrocochleography (ECochG) in subjects with auditory neuropathy spectrum disorder (ANSD)—who exhibit preserved outer hair cell (OHC) function—we captured cochlear microphonic (CM) tuning curves across frequencies of 500–4000 Hz. Our data demonstrate that as stimulus intensity increases, the peak of cochlear excitation shifts consistently toward more basal regions (Fig. 2); thus, at a given cochlear location, optimal responses occur at lower frequencies with rising intensities. In some cases, this frequency downward shift spanned up to an octave as sound levels increased from 36 dB SPL to 106 dB SPL (Fig. 5). Additionally, high-intensity stimuli produced broader spatial excitation patterns, whereas near-threshold stimuli yielded more localized responses. These findings align closely with earlier animal-model observations demonstrating intensity-dependent shifts in auditory nerve fiber firing (*29, 30*) and outer hair cell responses (*16*). Notably, the ECochG-derived best-frequency maps at high intensities (∼83 dB SPL) did not significantly differ between subjects with ANSD and those with adult-onset post-lingual hearing loss from our previous work (*21, 22*) (Fig. 3H), suggesting that cochlear frequency-place coding converges across these groups at elevated sound levels. This similarity likely results from saturation of active cochlear mechanisms mediated by OHCs at high intensities, causing passive cochlear mechanics to dominate, thereby producing consistent basalward shifts and broader excitation independent of the underlying hearing loss etiology (*16, 31, 32*).

Current CI programming uses a fixed, logarithmic frequency-to-electrode map that neither adjusts for stimulus intensity variations nor is grounded in direct physiological evidence. Thus, there is inherent place-frequency mismatch, partly stemming from the absence of a physiologically-derived target and the lack of attention to intensity-based shifts and spread of excitation. In our previous work, we showed that frequency-place mismatch, based on ECochG measures, better correlated with speech perception (i.e., CNC word scores) at 6 months than those derived from Greenwood and/or Stakhovskaya et al. functions (*22*). Our findings in the current work further reveals the importance of using a frequency-position function that is intensity-dependent, a factor omitted from previous models. Aligning mapping strategies more closely with natural cochlear responses might be expected to improve outcomes. However, whether an intensity-adjusted map will ultimately benefit CI users remains unknown. Some studies have reported advantages with anatomical fitting (*33–39*) although others have found no benefit—or even poorer performance— compared to standard care (*40–43*). It stands to reason that this lack of clear benefit relates to the choice of Greenwood’s map as an inappropriate target. Applying Greenwood’s equation to current CI electrode configurations presents further challenges, such as the potential loss of low-frequency information when deactivating basal electrodes that correspond to anatomical frequencies beyond the device’s upper limit. For patients who do not fully adapt to standard mappings or for bimodal CI users (e.g., those with single-sided deafness), reducing frequency-place mismatch provides a rational approach to improving performance.

Another novel aspect of our study is the demonstration that the spatial spread of acoustically evoked responses varies with stimulus intensity (Fig. 6A**-E**). Despite the limited number of electrode contacts (n = 22) in our array, high-intensity stimuli consistently elicited a broad excitation pattern, while near-threshold levels activated only localized regions. Both the extent of excitation and the accompanying shifts in BF location are critical for accurately modeling cochlear mechanics in CI programming.

Sagi et al. (2025) recently proposed an intensity-adjusted cochlear frequency-to-place map based on animal data, which corrects the standard Greenwood function by accounting for intensity-dependent shifts in BF location (*26*). Their model indicates that at moderate intensities, such as 70 dB SPL, the BF shifts basalward relative to the characteristic frequency for cochlear locations up to 600° from the base, while apical regions exhibit apical shifts, reflecting the dynamic influence of cochlear mechanics across intensity levels. In our study, the ECochG-derived map at near-threshold intensities (43 dB SPL) closely aligns with Sagi’s intensity-adjusted function (Fig. 3F). However, at 70 dB SPL, our human-derived map demonstrates a more pronounced basal shift compared to Sagi’s estimation (Fig. 3G). This discrepancy may stem from species differences in cochlear anatomy and physiology, as well as the unique profile of our subjects with ANSD, who exhibit preserved outer hair cell function based on ABR and OAEs, yet uncharacterized cochlear mechanics to date. Methodological variations—such as our use of peak CM response amplitudes versus Sagi’s centroid-based BF calculations—could also contribute to this difference. Our findings both corroborate and extend Sagi’s work by revealing significant tonotopic shifts across a broad intensity range (33–103 dB SPL) in humans, accompanied by broader spatial spread of excitation at higher intensities. This highlights the inadequacy of static, single-intensity maps like Greenwood’s for capturing the dynamic nature of cochlear frequency coding. Consequently, future iterations of CI stimulation paradigms could incorporate intensity-dependent adjustments to optimize frequency allocation and better replicate natural cochlear responses, potentially improving patient outcomes. However, implementing such strategies may require advancements in current CI hardware and software to accommodate these dynamic strategies.

A limitation of our methodology is that it provides an indirect approximation of the cochlear traveling wave by relying primarily on the peak electrical response of the CM. Although this peak may not precisely coincide with the true mechanical peak, animal studies have demonstrated a strong coupling between electrical and mechanical cochlear activity (*44, 45*), supporting the validity of our approach. Additionally, the limited number of electrodes and restricted apical insertion depth constrain our ability to capture the full extent of excitation, particularly in inter-electrode and deep apical regions. At low intensities—especially for 500 Hz—the estimated response may be shifted too apically, extending beyond the most apical electrode of the implant, which further limits the accuracy of the frequency-position estimation in this region. Other limitations include a modest sample size, incomplete characterization of hearing loss etiologies, reliance on pure-tone stimuli, and the variability inherent to intraoperative recordings. Finally, the study does not correlate these physiological measures with long-term perceptual or clinical outcomes, leaving the functional implications for CI performance to be determined.

Anatomically based place-frequency functions have traditionally relied on Greenwood’s human coefficients (1961, 1990) to relate cochlear position to CF, despite limited direct human physiological data. Animal studies have long reported that the peak of the cochlear response—the BF location—shifts basally away from the CF at higher (i.e. suprathreshold) stimulation levels. In this study, recordings from CI subjects with preserved outer hair cell function enabled us to develop an intensity-dependent place-frequency function in humans. High-intensity stimulation (>80 dB SPL) shifts BF locations in a basal (up to 158°) or frequency-downward (∼1 octave) direction relative to near-threshold levels. Higher intensities also produced broader spectral spread of excitation, whereas near-threshold levels result in a more localized response. These findings suggest that a static, frequency-fixed map cannot replicate the dynamic range of normal cochlear responses. By incorporating intensity-dependent adjustments, our proposed function offers a more physiologically accurate reference for electrode frequency assignments, with the potential to optimize cochlear implant programming and improve auditory outcomes for CI users.

## Materials and Methods

### Study Design

This was a prospective, observational study designed to investigate intensity-driven shifts in tonotopic coding in the human cochlea using intracochlear electrocochleography (ECochG). The study was conducted at Washington University in St. Louis with IRB approval (IRB #202007087). Five participants (median age 18 months, range 10 months – 8 years; 3 female, 2 male) were enrolled based on availability and meeting inclusion criteria, with three undergoing bilateral cochlear implantation, resulting in eight ears tested. The sample size was determined by the number of patients undergoing cochlear implantation during the study period who met the inclusion criteria, as this is a rare and specific population. Data collection was stopped once all eligible participants who underwent surgery during the study period had their data collected and analyzed. Exclusion criteria included middle ear pathology, revision cochlear implantation surgery, or lack of a patent external auditory canal, as the study required air-conducted acoustic stimulation. Subjects were further excluded if no clear tonotopic ECochG pattern emerged following electrode pull-back, as previously described (*46*).

### Cochlear Implant Surgical Procedure

All cochlear implant surgeries were performed by the senior author using a consistent surgical approach. A standard mastoidectomy and facial recess procedure was conducted to access the cochlea. The round window niche overhang was partially removed to facilitate electrode insertion. Depending on the orientation of the round window membrane, the electrode array was inserted either directly through a round window incision or after creating a marginal cochlear opening. A perimodiolar electrode array (Model CI632; Cochlear Corp., Sydney, NSW, Australia) was used for all insertions. Intraoperative radiographs confirmed the correct coiling of the electrode array within the cochlea. To prevent perilymph leakage, the cochleostomy was sealed with temporalis muscle or fascia following array placement. In one subject undergoing a translabyrinthine craniotomy for acoustic neuroma resection, the cochlear implant was removed at the end of the procedure due to non-preservation of the cochlear nerve. The receiver-stimulator unit was positioned securely in a subperiosteal pocket.

### Intracochlear Electrophysiological Measurements

Prior to surgical site sterilization, an ER3-14A insert earphone (Etymotic, Elk Grove Village, IL, USA) was placed in the external auditory canal. After implanting the electrode array into the cochlea, a telemetry coil was positioned over the skin and aligned with the cochlear implant antenna using a sterile ultrasound drape. To ensure accurate and reliable measurements, all 22 electrodes within the array were conditioned with reference to the case ground, minimizing electrical noise and establishing a common reference potential. Tone burst stimuli were delivered at frequencies of 250, 500, 1000, 2000, 3000, and 4000 Hz, each presented in both condensation and rarefaction phases, with a minimum of 30 repetitions per phase. The stimulus intensities for these frequencies (250-4000 Hz) were set at 114, 102, 98, 106, 104, and 107 dB SPL, respectively, based on the maximum output capacity of the speaker. Each tone burst had a duration of 14 ms, including a 1 ms rise and fall time shaped by a Blackman window, with a recording epoch of 18 ms starting 1 ms prior to stimulus onset. Recordings were sampled at a rate of 20 kHz, capturing electrophysiological responses across all 22 electrodes in the array. To assess the effect of intensity on the tonotopic map, the stimulus intensity was gradually reduced in 10-15 dB increments until a shift in the tonotopic response was detected. This process continued until either a further shift in the map was observed or the response was no longer detectable.

### Electrophysiological Signal Analysis

The recorded electrophysiological responses, collected as separate condensation and rarefaction phases, were processed offline using custom software in MATLAB R2023a (MathWorks Corp., Natick, MA, USA). To enhance signal analysis, we generated a difference curve by subtracting the rarefaction phase from the condensation phase. From this difference curve, we isolated the ongoing response segment for fast Fourier transformation (FFT), which allowed us to determine response amplitudes for each stimulus frequency. Cochlear microphonic (CM) tuning curves were then constructed across the entire electrode array using these frequency-specific response amplitudes. Significant responses were defined as those where the signal magnitude exceeded the noise floor by three standard deviations, consistent with established criteria.(*21, 22, 47–49*) For these recordings, the noise floor was approximately 0.3 μV. The “best frequency” (BF) location was identified as the electrode along the array producing the highest response amplitude to a given stimulus, indicating its correspondence to a specific location on the basilar membrane.

To quantify the spectral spread as a function of stimulus intensity, we examined the number of electrodes exhibiting measurable responses for each pure tone frequency stimulus. Given that the electrode array is limited to 22 evenly spaced contacts and has a maximal, angular insertion depth (∼425°), this approach has inherent constraints. To account for these limitations, we fit the data using a linear mixed effects model with a quadratic term to capture non-linear effects in the spread of excitation.

### Determination of Electrode Position

Electrode angular insertion depth (AID) was measured using intraoperative radiography combined with a validated three-dimensional helical scala tympani model.(*50*) This approach was selected to avoid the unnecessary radiation exposure associated with CT imaging in pediatric patients. The model relies on equations derived from a study of 30 CI recipients (*51*). During the procedure, an intraoperative X-ray was obtained, and the helical model was adjusted to match the visible cochlear anatomy by precisely aligning with the electrode array’s trajectory. This adjustment involved scaling, rotating, and positioning the model to accurately trace the implanted electrode’s path. Validation studies have demonstrated that this method estimates AID within 10 degrees of values obtained from CT imaging, confirming its reliability and accuracy for clinical use.

### Tonotopic Map in a Normal Hearing Subject

An additional subject with normal audiometric thresholds and word recognition scores underwent a translabyrinthine craniotomy for vestibular schwannoma removal, intentionally sacrificing residual hearing. Prior to labyrinthine resection, cochlear implantation and electrophysiological measurements proceeded as described above. BF locations across the electrode array were recorded at maximum stimulus intensities for frequencies between 250 and 4000 Hz. Subsequently, stimulus intensity was systematically reduced toward auditory threshold, and CM tuning curves were regenerated to characterize intensity-dependent shifts in cochlear tonotopic organization.

### Statistical Analysis

Statistical analyses were performed using MATLAB R2023a (MathWorks Corp., Natick, MA, USA). The primary outcome measures were the AID of the BF electrode and the number of activated electrodes as a function of stimulus intensity. To assess intensity-dependent shifts in the BF electrode position, a log-linear mixed-effects model was employed for each frequency (500, 1000, 2000, 3000, and 4000 Hz), using stimulus intensity (dB SPL, log-transformed) as a fixed effect and subject as a random intercept to account for repeated measures within subjects. Adjusted R² values were computed to evaluate the goodness-of-fit for each model, and p-values were obtained to assess the significance of the intensity effect. To compare ECochG-derived BF locations against established frequency-position maps (Greenwood and Sagi), Wilcoxon signed-rank tests were conducted due to the nonparametric distribution of the angular insertion depth data.

Differences between ECochG-derived maps at various intensities (43, 58, and 83 dB SPL) and the corresponding frequency-place maps were assessed, and statistical significance was defined at p < 0.05. To evaluate differences in ECochG-derived BF maps between subjects with ANSD and those with adult-onset hearing loss, a Wilcoxon signed-rank test was similarly applied. For the analysis of spatial excitation spread, a quadratic mixed-effects model was fit to examine the number of activated electrodes as a function of stimulus intensity, with both linear and quadratic intensity terms included as fixed effects and subject as a random intercept. Significance of the linear and quadratic terms was assessed, and confidence intervals (95%) were calculated to quantify uncertainty. All results were reported with estimates, standard errors (SE), confidence intervals (CI), and p-values. A p-value less than 0.05 was considered statistically significant.

## Data Availability

All data produced in the present study are available upon reasonable request to the authors.

## Acknowledgments

Cochlear Corp provided equipment to measure real-time electrocochleography responses.

## Author contributions

Conceptualization: CAB, AW

Methodology: CAB, AW, MAS, SML, AJO, PI, MW, JAV, JAH

Investigation: AW, MAS, SML, JAV, MW, PI Visualization: AW, MAS, SML, CAB Funding acquisition: CAB, AW, MAS, JAH Project administration: CAB, AW Supervision: CAB

Writing – original draft: AW, CAB

Writing – review & editing: AW, CAB, MAS, SML, AJO, PI, MW, JAV, JAH

## Data and materials availability

All data associated with this study are in the paper and/or the Supplementary Materials.

